# U-shaped Relationship Between Body Mass Index and Intracerebral Hemorrhage-Related Functional Decline

**DOI:** 10.1101/2024.07.01.24309789

**Authors:** Yuji Kanejima, Masato Ogawa, Kodai Ishihara, Naofumi Yoshida, Michikazu Nakai, Koshiro Kanaoka, Yoko Sumita, Takuo Emoto, Yoshitada Sakai, Yoshitaka Iwanaga, Yoshihiro Miyamoto, Tomoya Yamashita, Kenichi Hirata, Kazuhiro P. Izawa

## Abstract

**BACKGROUND:** Intracerebral hemorrhage (ICH) has a high mortality rate, and even if patients survive, they are likely to have severe disability. Body mass index (BMI) is associated with ICH risk, and extremely low and high BMIs are associated with the site of ICH, which affects functional decline. However, few reports exist on ICH-related functional decline and BMI. This study aimed to clarify the relationship between BMI and stroke-related disability of patients with ICH.

**METHODS:** Patients with ICH registered in the Japanese Registry Of All Cardiac and Vascular Diseases Diagnosis Procedure Combination (JROAD-DPC) database from April 2016 to March 2020 were included. BMI was defined according to the World Health Organization Asia-Pacific classification. Functional disability was assessed using the modified Rankin Scale (mRS). ICH-related functional decline was defined as an increase in mRS score at discharge compared with that of the pre-stroke assessment.

**RESULTS:** This study included 155,211 patients with ICH whose median age was 72.0 years and mean BMI was 22.3 kg/m^2^. The ratio of patients with ICH who experience functional decline was 74.9%. The spline curve between BMI and ICH-related functional decline was U-shaped, revealing that the Normal to Obese I BMI groups (BMI: 22.2–30.4 kg/m^2^) exhibited reduced odds ratios for ICH-related functional decline. Hospitalization cost and BMI showed similar U-shaped patterns, with a BMI of 25.0 kg/m^2^ as the lowest point, regardless of age group.

**CONCLUSION:** In patients with ICH, those with both extremely low and high BMIs were more likely to experience functional decline after ICH onset, which resulted in increased hospitalization costs. To reduce ICH-related functional decline, patients should be managed at a normal to slightly obese BMI.

## INTRODUCTION

### Epidemiology of Intracerebral Hemorrhage

Stroke is the second leading cause of death worldwide, with 12 million strokes and 101 million prevalent cases in 2019.^1^ Over the past 30 years, the numbers of onsets of stroke, deaths, and prevalent cases have increased around the world,^1^ making the investigation of effective strategies against stroke an important issue. Intracerebral hemorrhage (ICH) is the second most common form of stroke (27.9%).^2^ The mortality rate for ICH is higher than that for ischemic stroke, and even if patients survive, they are more likely to have severe disability.^2^ In fact, only 35% of patients with ICH had a favorable physical outcome at 30 days after onset,^3^ and only 41% of patients who experienced ICH were independent in activities of daily living (ADL) at one year after ICH onset.^4^ Disability-adjusted life-years due to stroke are also increasing.^1^ Improvement of stroke-related disability and reintegration into society after ICH are also important issues.

### Relationship Between ICH and Body Mass Index

Body mass index (BMI) has been associated with ICH risk.^5,6^ In general, obesity causes increased mortality or morbidity of cardiovascular disease in patients with ischemic stroke and ICH.^7–9^ ^10^ However, several reports have shown that the obesity paradox leads to lower mortality in overweight and obese patients with ICH.^8,11,12^ Whether obesity is advantageous for patients with ICH remains controversial.

The American Heart Association supposed the ideal BMI to be <25 kg/m^2^.^10^ However, there are regional differences in BMI, and the Asian population tends to have a lower BMI.^13,14^ In Japanese people, a BMI of about 22 kg/m^2^ is associated with the lowest incidence of obesity-related diseases, and the Japan Society for the Study of Obesity defined the standard weight for Japanese people as weight equivalent to a BMI of 22 kg/m^2^.^15^ Extremely low BMI (<18.5 kg/m^2^) and high BMI (>30.0 kg/m^2^) are associated with the site of ICH.^16,17^ Symptoms and degree of functional impairment vary depending on the site of hemorrhage.^18,19^ It remains unclear what range of BMI results in the fewest stroke-related symptoms in Japanese patients with ICH.

### Objective

In patients with ischemic stroke, BMI has been reported to be associated with stroke-related disability,^8^ and although ICH is more likely to cause severe disability,^2^ the relationship between BMI and stroke-related disability of patients with ICH is poorly documented and unclear. In the present study, we hypothesized that patients with higher BMI would likely experience functional decline after ICH. The study aim was thus to clarify the relationship between BMI and functional decline in Japanese patients with ICH. As a secondary outcome, we also examined the relationship between BMI and hospitalization costs in patients with ICH.

## METHODS

### Study Design

This was a retrospective cohort study based on the Japanese Registry Of All Cardiac and Vascular Diseases Diagnosis Procedure Combination (JROAD-DPC) database. The JROAD-DPC database targets data on cardiovascular or cerebrovascular disease patients treated in facilities affiliated with the Japanese Circulation Society.^20^ The JROAD-DPC database contains the following data: unique hospital identifier, patient age, sex, primary diagnoses, comorbidities, drugs, devices, diagnostic and therapeutic procedures, length of stay, and discharge status. The International Statistical Classification of Diseases and Related Health Problems 10th Revision (ICD-10) was used for primary diagnosis and comorbidity in the JROAD-DPC database.^20^ The study was carried out in accordance with the Declaration of Helsinki ^21^ and was approved by the Kobe University Institutional Review Board (Approval No. B210052).

The inclusion criterion was hospitalization for ICH from April 2016 to March 2020. Exclusion criteria were (1) age <20 years old, (2) length of hospital stay <3 days, (3) no emergency admission, (4) hospitals with <10 beds, (5) hospital death, and (6) no brain rehabilitation during hospitalization. The ICD-10 divided ICH into six groups: subcortical (I610), cortex (I611), brainstem (I613), cerebellum (I614), intraventricular (I615), and other types.

### Variables

We collected data on the following variables: age, sex, hospital code, hospital scale, hospitalization days, hospitalization cost, disposition, comorbidities (hypertension, diabetes mellitus, dyslipidemia, congestive heart failure, stroke, Charlson Comorbidity Index score),^22^ smoking, treatment (drainage, evacuation, shunt), complications (pneumonia, urinary tract infection [UTI], bedsores), level of ADL at admission and discharge, and pre-stroke modified Rankin Scale (mRS) score and mRS score at discharge. The drainage, evacuation, and shunt treatment data were collected using the billing codes in reception data. ADL were assessed with the Barthel index,^23^ which was done immediately after admission. We defined hospital scale as small: <100, medium: 100–500, and large: >500 beds. Hospitalization costs were converted into US dollars based on the exchange rate on January 10, 2024 (one US$=145 Japanese yen).

### BMI and Classification

BMI was calculated by dividing body weight by the square of the height (kg/m^2^). According to the World Health Organization (WHO) Asia-Pacific classification,^13^ we divided participants into five BMI groups at admission, as follows: Underweight (BMI ≤ 18.5 kg/m^2^), Normal (18.5 kg/m^2^ < BMI ≤ 23 kg/m^2^), Overweight (23 kg/m^2^ < BMI ≤ 25 kg/m^2^), Obese I (25 kg/m^2^ < BMI ≤ 30 kg/m^2^), and Obese II (30 kg/m^2^ ≤ BMI).

### Modified Rankin Scale

The primary outcome was ICH-related functional decline.^24^ The mRS is the most frequently used assessment tool for stroke-related disability^25,26^ as a measure of outcome in stroke studies.^27^ The mRS score ranges from 0 (no symptoms at all) to 6 (dead), with a higher score indicating increased disability. In the JROAD-DPC database, pre-stroke mRS was assessed for stroke-related disability before the onset of stroke,^28,29^ and the mRS was used again for patient assessment just before discharge. If the mRS score at discharge was higher than the pre-stroke mRS score, we regarded this as indicative of ICH-related functional decline.

### Statistical Methods

Statistical analysis was conducted using the software R version 4.3.2.^30^ Continuous variables are expressed as median [interquartile range] and categorical variables as number (%).

Initially, a comparative analysis between the BMI groups by Asia WHO classification was conducted with ANOVA for continuous variables and by chi-square test or Fisher’s exact test for categorical variables and included missing values for several variables. Next, multiple mixed-effect logistic regression analysis was conducted for ICH-related functional decline as the dependent variable with the hospital code as a random intercept. Independent variables in the developed model were as follows: age, sex, BMI, hospital scale, smoking, ICH type, hypertension, diabetes mellitus, dyslipidemia, congestive heart failure, stroke, treatment (drainage, evacuation, shunt), complication (pneumonia, UTI, bedsores), and pre-stroke mRS score. We excluded missing values of the model variables and a pre-stroke mRS score of 5 as a further increase in mRS score to 6 would mean hospital death. The odds ratio (OR) of the outcome referenced normal BMI, large hospital scale, subcortical ICH, and pre-stroke mRS score=0. Finally, we created restricted cubic spline models based on the logistic regression analysis results for ICH-related functional decline. The spline models involved explanatory variables of the model. In addition, spline models were also created with hospitalization costs as the objective variable for patient ages 75 or over and under 75 years. The explanatory variables were based on the model.

## RESULTS

### Overall Trends

There were 222,318 patients with ICH admitted to the affiliated hospitals from April 2016 to March 2020. Among these patients, 67,107 who met the exclusion criteria as shown in Figure 1 were excluded. Thus, the final analysis included 155,211 patients with ICH.

**Figure 1.**
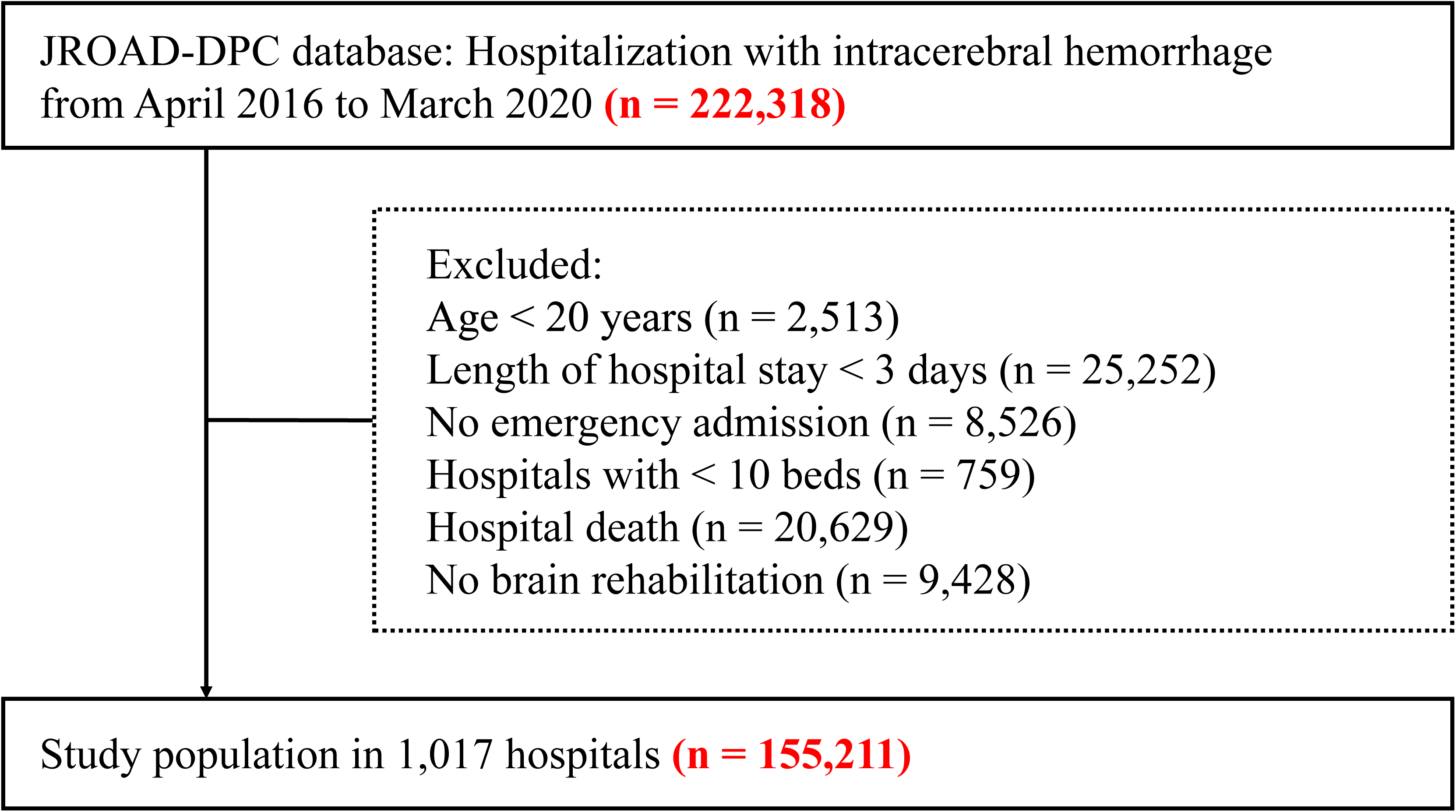
Flow of the present study. This flowchart depicts selection of the present study patients with ICH using the JROAD-DPC database. JROAD-DPC, Japanese Registry of All Cardiovascular Diseases Diagnosis Procedure Combination.

Overall, the median age was 72.0 [61.0, 81.0] years old, the female ratio was 44.8%, and the mean BMI was 22.3 [19.8, 25.1] kg/m^2^. Among the BMI groups, Normal comprised the greatest ratio of patients (38.1%), followed by Obese I (17.5%), Overweight (15.9%), Underweight (13.2%), and Obese II (5.3%). The ICH types included subcortical (78.1%), cortex (1.2%), brainstem (5.7%), cerebellum (8.6%), and intraventricular (2.4%). ICH-related functional decline occurred in 74.9% of the patients with ICH. The percentage of patients with a pre-stroke mRS of 0 was the highest at 56.5%, and that of patients with a mRS of 4 at discharge was highest at 32.4%. The median number of hospitalization days was 26.0 [17.0, 39.0], and hospitalization costs were $8800 [$5879, $13,929] (Table 1).

### Comparison by BMI

Lower BMI was associated with higher age and female ratio. In addition, patients with lower BMI had a longer number of hospitalization days, higher hospitalization costs, and a lower rate of home discharge. Among ICH types, subcortical ICH remained the most common in all BMI groups, and brainstem ICH was more common in patients with a higher BMI. Coronary risk factors such as hypertension, diabetes mellitus, dyslipidemia, and smoking were more prevalent at a higher BMI, whereas complications such as pneumonia, UTI, and bedsores occurred more frequently in patients with a lower BMI. A pre-stroke mRS score of 0 was the most common in each BMI group (42.1–70.8%). The rates of ICH-related functional decline for each BMI group were Underweight (70.4%), Normal (73.8%), Overweight (75.0%), Obese I (75.8%), and Obese II (76.3%). Patients with lower BMI had a higher percentage of mRS score of 5 (e.g., Underweight: 31.0%, Obese II: 13.6%) at discharge.

### Odds Ratio of ICH-Related Functional Decline

The multiple mixed-effects logistic regression analysis was conducted on patients with complete data (n=115,179). The analysis revealed that the OR of ICH-related functional decline was lower in the Overweight and Obese I groups and higher in the Underweight and Obese II groups, with the Normal group as a reference (Table 2). The OR of ICH-related functional decline was lowest in the Overweight group (OR: 0.90, 95% CI: 0.85–0.94). ICH-related functional decline was likely to be observed in patients with subcortical ICH, some treatments (drainage, evacuation, shunt), and complications (pneumonia, UTI, bedsores). The adjusted R^2^ value of this multiple regression model was 0.48. The spline curve for BMI and the OR of ICH-related functional decline was U-shaped, with BMI 25.4 kg/m^2^ as the lowest point (Figure 2). The range of BMI of 22.2–30.4 kg/m^2^ had an OR of <1 for ICH-related functional decline. Outside of this BMI range, the OR for a declining mRS tended to increase in patients with a lower BMI (Underweight).

**Figure 2.**
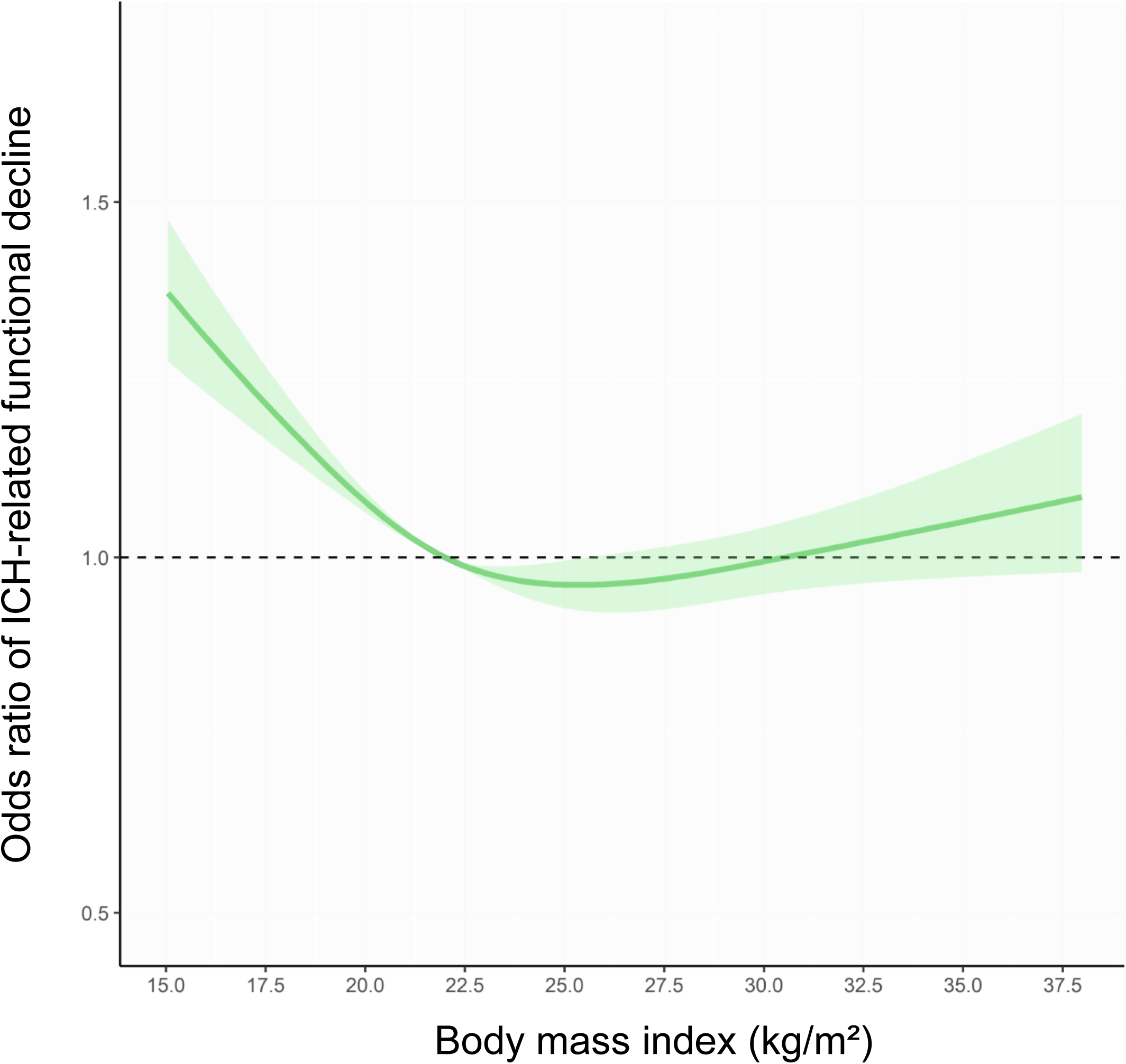
BMI spline curve for intracerebral hemorrhage-related functional decline. This spline curve was created with body mass index on the x-axis and odds ratio of ICH-related functional decline on the y-axis. ICH, intracerebral hemorrhage.

### Hospitalization Costs

The spline curves for BMI and hospitalization costs revealed that individuals aged 75 years or older incurred higher hospitalization costs than the overall median of $8,801. The curves exhibited U-shapes for both age groups of patients: those 75 years or older and those under 75 years. Notably, both curves reached their lowest points at a BMI of 25.0 kg/m^2^ (Figure 3).

**Figure 3.**
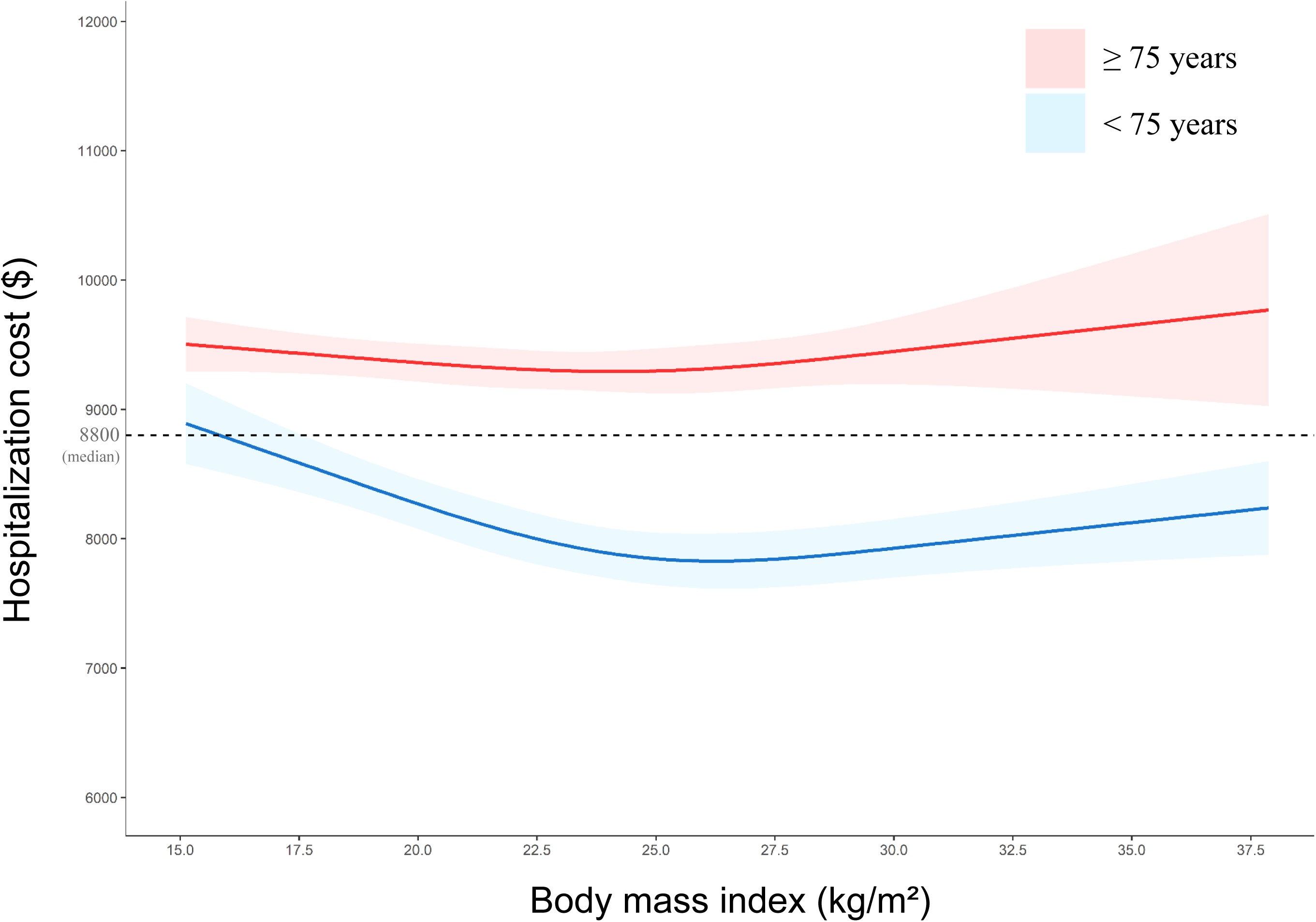
BMI spline curves for hospitalization cost. Spline curves were stratified by age under 75 (blue) or age 75 or over (red), with body mass index on the x-axis and hospitalization cost on the y-axis.

## DISCUSSION

### Key Results

The present study included 155,211 patients with ICH. The distribution of ICH types was as follows: subcortical (78.1%), cortex (1.2%), brainstem (5.7%), cerebellum (8.6%), and intraventricular (2.4%). ICH-related functional decline occurred in 74.9% of the patients with ICH. Multiple mixed-effects logistic regression analysis revealed that the OR of ICH-related functional decline was lower in the Overweight and Obese I groups. The spline curve for the OR of ICH-related functional decline exhibited a U-shape, with a BMI of 25.4 kg/m^2^ as the lowest point. The BMI range of 22.2–30.4 kg/m^2^ had an OR of <1 for ICH-related functional decline. These results differed from our hypothesis that patients with a higher BMI would experience ICH-related functional decline. Additionally, the spline curve for hospitalization costs displayed U-shapes with BMI 25.0 kg/m^2^ at the lowest point for both age groups of 75 years or over and under 75 years.

### Generalizability

Compared to a Japanese DPC study that did not include data from the JROAD-DPC database, the distribution of BMI followed a trend similar to that observed in the present study.^31^ We also confirmed that patients in the higher BMI group tended to be younger, more were male, had a higher smoking prevalence, and were more likely to have hypertension and diabetes mellitus.^31^ Despite the low home discharge rate of 26.9% in the present study, a nationwide U.S. database study of inpatients with ICH similarly reported a home discharge rate of approximately 20%.^32^ Therefore, when considering patients with ICH, their characteristics do not differ significantly from those observed in other database studies.^31,32^

Compared to the Japan Stroke Data Bank report (n=9,741), the present study enrolled more patients with ICH (n=155,211),^33^ and although the age of patients in the present study was slightly older, the ratio of woman was not significantly different. Lobar ICH, such as subcortical ICH, was greater than that in previous studies, and a similar trend of patients with higher BMI being likely to have more lobar ICH was also observed. As in the previous study,^33^ the mRS at discharge was higher in the present study groups with extremely low and high BMI. The present study supported the findings of these previous studies, with the additional finding that BMI was associated with ICH-related functional decline in a larger number of ICH cases.

In the present study, a pre-stroke mRS score of 0 was the most common (56.5%), which aligns with a generally similar trend observed in a previous study showing that pre-stroke mRS score of 0 accounted for 51% of the total.^34^ The pre-stroke mRS score serves as a valuable tool for clinicians in treatment planning and delivery.^34^ At discharge, an mRS score of 4 (moderately severe disability) was the most common. Another study also reported the same score as being the most frequent mRS score at discharge, except for the mRS score of 6 (death). Over the one-year follow-up period, 34% of all patients showed functional improvement and shifted to mRS scores of 0–3.^35^ While many patients with ICH experience significant improvement, often from very severe disability, during the first three months, functional progress across the disability spectrum occurs from 3 to 6 months after the onset.^35^ The mean length of hospitalization in the present study was 26.0 days, with limited follow-up carried out after the onset of ICH. Although post-discharge follow-up data were unavailable in this study, continuing rehabilitation and other interventions to enhance function after discharge remain crucial.

### Interpretation

The present study revealed that individuals with a slightly higher BMI (ranging from 22.2 to 30.4 kg/m^2^) had a lower OR for functional decline in the patients with ICH, similar to the concept of the obesity paradox observed in mortality studies.^36^ In stroke patients, stress-related neuroendocrine and autonomic activation, inflammatory cytokines, increased oxygen-free radical load, and systemic hormonal changes collectively contribute to an overall catabolic state.^37^ Obese patients may be better equipped to tolerate such nutritional disturbances due to greater fat and muscle mass metabolic reserve.^12^ Additionally, another potential mechanism underlying the obesity paradox is resistance to unintended weight loss after ICH.^12^ Patients with ICH are more susceptible to long-term weight loss compared to patients with ischemic stroke.^38^ Conversely, low BMI negatively impacts the resumption of oral intake in stroke patients. ICH patients with low BMI may experience insufficient nutrient intake, leading to further weight loss and hypercatabolism.^39^ We speculate that with their greater metabolic reserve, overweight to obese patients may possess a physiological advantage against functional decline as indicated by the mRS.

Intraventricular ICH is more likely to result in functional disability after discharge. Conversely, subcortical and cerebellar ICH have exhibited an OR of 0.26 for disability.^3,4^ Therefore, the impact of different ICH sites on functional impairment is significant. ICH can be categorized into lobar ICH (isolated to the cortex) and deep ICH (such as in the brainstem). Extreme low or high BMI has been associated with an increased risk of deep ICH.^16,17^ In our study, cerebellar hemorrhage was more common in the Normal and Overweight groups, and the OR for ICH-related functional decline was lower in these groups. Conversely, the other BMI groups were associated with deeper ICH, including intraventricular and brainstem hemorrhages. Therefore, depending on the BMI at onset, trends may emerge regarding the site of ICH and the degree of functional disability. The BMI of patients should be managed to ensure that they maintain a BMI within the Normal to Obese I WHO categories and not those of the Underweight or Obese II categories..

The overall median cost of hospitalization of ICH patients in this study was $8,800, which aligns with findings from a previous Japanese study reporting $8,301.^40^ The spline curves indicate that hospitalization costs were $500 to $2,000 higher for individuals aged 75 or over than for those under 75 years, regardless of BMI. Additionally, the spline curve exhibits a U-shape, similar to that for ICH-related functional decline, suggesting that preventing functional decline in patients with ICH may lead to reduced hospitalization costs.

### Strengths and Limitations

The strength of the present study lies in it being the first, to our knowledge, to examine the transition of functional decline before and after the onset of ICH with a focus on BMI. The spline curves revealed that the Normal to Obese I groups at admission were associated with favorable outcomes regarding functional decline and hospitalization costs. However, the present study also has several limitations. First, the JROAD-DPC database includes only facilities participating in the Japan Circulation Society (JCS). The number of facilities treating cerebrovascular disease that have joined the JCS is less than that of the facilities for treating heart disease. Thus, whether the present study reflects the real world in Japan requires consideration. Second, ICH and comorbidities were evaluated using the ICD-10, which may have latent selection bias. In addition, we could not consider the presence of ICH histories nor the amount and extent of hemorrhage in ICH, which affects the prognosis. Third, rehabilitation is a major part of patient care, and its intervention significantly impacts functional transition after the onset of stroke.^41^ Therefore, consideration should be given to the fact that the present study was limited to only those patients who underwent rehabilitation, which could affect the primary outcome. Finally, stroke-related disability was assessed by the mRS. However, the mRS has been criticized for its poor reproducibility, score grades that are not proportional to each other, its emphasis on motor function rather than cognitive or social function, and less consideration of other symptoms in neurological severity, such as state of consciousness and sensory function.^26^ In addition, as we could not use the pre-stroke mRS to assess the patients’ pre-stroke disability directly, the assessments were based on the collection of information on functional status and living conditions of the patients before the onset of ICH. Although the present study used the JROAD-DPC database, further research is needed using detailed data on ICH in clinical practice.

## CONCLUSION

This study represents the first examination of the relationship between BMI and stroke-related disability (measured by the mRS) following the onset of ICH. The final analysis included 155,211 patients with ICH, of whom 74.9% experienced functional decline.

Notably, unlike our hypothesis, the analysis revealed that individuals in the Normal to Obese I BMI groups (with a BMI range of 22.2–30.4 kg/m^2^) exhibited reduced ORs for ICH-related functional decline. Furthermore, the spline curves showed that hospitalization costs followed U-shaped patterns, with a BMI of 25.0 kg/m^2^ as the lowest point, regardless of age group.

Similar to findings in mortality studies, the obesity paradox concept may also apply to functional decline after ICH, underscoring the importance of appropriate BMI management of patients at risk for stroke.

## Non-standard Abbreviations and Acronyms

ADL: activities of daily living
BMI: body mass index
ICD-10: International Statistical Classification of Diseases and Related Health Problems 10th Revision
ICH: intracerebral hemorrhage
JROAD-DPC: Japanese Registry Of All Cardiac and Vascular Diseases Diagnosis Procedure Combination
mRS: modified Rankin Scale
UTI: urinary tract infection
WHO: World Health Organization

## Acknowledgment

We appreciate the contributions of the investigators, clinical research coordinators, and data managers involved in the JROAD-DPC study.

## Data availability

Deidentified participant data in the JROAD-DPC study will not be shared.

